# Cross sectional study of knowledge, attitude and practice among general population towards COVID19 vaccines in Duhok province, Kurdistan region of Iraq

**DOI:** 10.1101/2023.04.01.23288042

**Authors:** Ramis Imad Elyas, Halima Adil Abdulrahman, Rozan Sagvan Ismaeel

## Abstract

**Background:** Vaccines are immunization against diseases and leads to saving millions of lives every year. However, after the availability of COVID-19 vaccines, little information is available on the public knowledge and attitudes towards the COVID-19 vaccines in Kurdistan-Iraq.

**Aim:** This study aimed to investigate the knowledge, attitude and practice toward the COVID-19 vaccines among general population at Duhok province, Kurdistan region, Iraq.

**Methods:** The cross-sectional study was done between November 1^st^, 2022 and march 1^st^, 2023 at Duhok province, Kurdistan region, Iraq including Duhok City, Zakho, Semel and surrounding area) toward COVID-19 vaccines. It included 759 randomly selected participants answering a structured questionnaire who were interviewed face-to-face by the authors. The participants ages ranged from 18 to 75 years. The survey questionnaire was divided into three parts, the first part was sociodemographic characteristics. The second part was composed of eight questions of knowledge regarding the COVID-19 vaccine and third part was 6 statements about Attitudes toward COVID-19 vaccines.

**Findings:** The mean age of the respondents was 32.95 years (SD±12) and more than half of them (52.3%) were males. About 55% of the respondents reported that they had infected with COVID-19. About 25.3% of the subjects were employed and only 18.3% had chronic diseases. Around 55% of the participants reported that they have previously infected with COVID-19. The majority of the participants (99.60%) had heard of COVID-19 vaccine, almost (68%) of the participants trusted COVID-19 vaccine and reported that the vaccine is safe. Almost three-quarters (74.04%) of the participants were strongly agreed that it is important to get a vaccine to protect the people from COVID-19. According to the survey results, a significant proportion of the participants, specifically 62.58%, believed that COVID-19 vaccines offer protection against the disease. It was notable that a high percentage of the participants, approximately 86.17%, were aware of the potential side effects associated with the vaccine. Moreover, an overwhelming majority of the participants, nearly 96.31%, were knowledgeable that the vaccination process would require two or more doses.

**Conclusions:** The history of chronic disease, source of vaccine knowledge, education level, occupation, and employment states were factors that affected the willingness to accept the vaccine. The most trusted sources of information on COVID-19 vaccines were social media. Therefore, the willingness to take the COVID-19 vaccine can be supported by utilizing social media and television to spread awareness about the vaccine’s safety and efficacy.

## Introduction

On December 31 the Wuhan municipal health commission faced a medical issue, some of the patients were identified to have ground-glass appearance on Imaging tests, decreased average white blood cells (lymphocyte), platelet count and abnormal renal and liver function (1) this was responded by Chinese center for disease control and prevention it also notified WHO, on January 7 2020 the causative agent recognized as novel coronavirus (2). SARA-COV-2 was known to be the cause behind COVID-19 pandemic, many of these coronaviruses can also infect animal species as well (3), and on 9^th^ January 2020 a novel coronavirus-2019-ncov was formally known as the cause of an outbreak of viral pneumonia in Wuhan city, Hubei province, China (4).

The outbreak has spread substantial to infect 19 other countries up to 31 January 2020. Then the infected disease has been named as coronavirus disease 2019 (COVID-19) by the World Health Organization (5) Most people infected with the virus suffered from mild to moderate respiratory illness and recover without requiring special treatment. However, some have been become seriously ill and require medical attention. Anyone can get sick with COVID-19 and become seriously ill or die at any age. The virus can spread from an infected person’s mouth or nose in small liquid particles when they cough, sneeze, speak, sing or breathe. The best way to prevent and slow down transmission is to be well informed about the disease, how the virus spreads, proper Protection and Get vaccinated (6). Globally, as of 8:47pm CET, 17 February 2023, there have been 756,581,850 confirmed cases of COVID-19, including 6,844,267 deaths, reported to WHO. As of 13 February 2023, a total of 13,195,832,385 vaccine doses have been administered (7,8)

This outbreak spread rapidly all over the world including Iraq (9-19) and till now researchers continue to do investigations as to its origin (20). The COVID-19 can lead to clinical features which are much similar to that of cold, flue, or pneumonia. The average duration between infection and symptom onset is between 7 to 14 days and of its common clinical features are fever and chills, headache, rhinorrhea, generalized body ache, nausea and vomiting. But patients may also develop loss of smell and taste and diarrhea. Most of the patients don’t need medical intervention though some may develop severe symptoms that require special treatment (21). In order to prevent and decrease the viral transmission is to have adequate knowledge about the disease and its routes of spread. Universally as of 4:24 pm CET 21^st^ February 2023, 757,264,511 cases of coronavirus were established which include 6,850,524 deaths. The best way to get protection is to receive vaccine and according to WHO as of 18^th^ February 2023 13,211,844,137 doses of vaccine are given to the population (22). Although no studies are conducted in Kurdistan region (23), this is the first study to investigate COVID-19 vaccination KAP, and associated sociodemographic characteristics among the general population of the Kurdistan region of Iraq but many studied conducted about KAP and medical education (24-28). and other infections including UTI, H. pylori and staph aureus among male and female[29-33]. Vaccination is a simple, safe, and effective way of protecting against harmful diseases, before come into contact with them. It uses your body’s natural defenses to build resistance to specific infections and makes your immune system stronger. However, because vaccines contain only killed or weakened forms of germs like viruses or bacteria, they do not cause the disease or put at risk of its complications.

The aims and findings of this study are expected to provide beneficial information to policymakers, about KAP toward COVID-19 vaccination among the Iraqi population. The findings may also inform public health officials on further public health interventions, awareness, and policy improvements pertaining to the COVID-19 outbreak. The finding of this study is also expected to provide useful information to policymakers, about KAP toward COVID-19 vaccination among the Iraqi population. The findings may also inform public health officials on further public health interventions, awareness, and policy improvements pertaining to the COVID-19 outbreak.

## Materials and methods

### 1. Study design

This population-based study was approved by Zakho medical University, Kurdistan Region, Iraq. A Cross sectional survey-based study was conducted between November 1^st^, 2022 and march1^st^, 2023. Sample approach adopted in this study where people from the different Kurdistan regions (Duhok, Zakho, Semel and surrounding area) were invited to participate. Prior to start of the study, the participants were told about the goal of the study and their consent was gained. A total of 759 randomly selected participants were recruited from the general population who were interviewed face-to-face by one of the authors. The participants’ ages ranged from 18 to 75 years (mean age: 32.95±12 SD). In total, there were 362 males and 397 females.

### 2. Study tool

The survey questionnaire was divided into three parts, the first part was about general questions that’s related to individual life (sociodemographic data) and health, including age, gender, marital status, education state, occupation, employment, smoking status, history of chronic illness, Infected with COVID19, Source about COVID-19 vaccination, Is the COVID-19 vaccine safe, Reasons for unwillingness to take the COVID-19 vaccine.

The second part was composed of eight questions of knowledge regarding the COVID-19 vaccine that answered by either yes or no such as, have you heard about the COVID-19 vaccine, the COVID-19 vaccine is safe with some side effects, the COVID-19 vaccine protects from getting COVID-19. And third part was 6 statements about Attitudes toward COVID-19 vaccines that answered by either agree, disagree or Neutral such as, it is important to get a vaccine to protect the people from COVID-19, Side effects will prevent me from taking a vaccine for the prevention of COVID-19, Pharmaceutical companies are going to develop safe and effective COVID-19 vaccines

### 3. Inclusion/exclusion criteria

Inclusion criteria including all individual live in Kurdistan Region, Iraq, if male or female above 18 years regardless of ethnicity, background, religion, occupation or socioeconomic state, the exclusion criteria were the individuals if under 18 years’ old or there is missing or incomplete data collection were all excluded.

### 4. Ethical approval

The survey was approved by ethics and scientific committee in university of zakho college of medicine, the ethical approval should be considered carefully because the participants have the right to know what is being done of their data and respect for individual rights including anonymity and confidentiality.

## 3. Results

### 3.1 Demographic characteristics of study participants

The study received a total of 759 participants from all over Kurdistan region. The mean age of the respondents was 32.95 years (SD±12) and more than half of them (52.3%) were males. Also, more than half of the respondents (56.91%) were married. About 30% are at primary level of education or less, and 37% had intermediate or secondary degree while higher educational level about 33%. And less than half (32%) with health-related educational backgrounds. Besides, 25.3% of the participants were employed and only 18.3% had chronic diseases. More than 70% of the participant were not smoker. About 55% of the respondents reported that they had infected with COVID-19. While about 45% not infected at all. almost (68%) of the participants trusted COVID-19 vaccine and reported that the vaccine is safe. As shown in table 1.

**Table 1.** Demographic details of study participants (n = 759).

|  |  |
| --- | --- |
| **Age (Year)** | n (%) |
| Mean ± SD | 32.95±12.08 |
| 18-25 years | 275 (36.23) |
| 26-35 years | 221 (29.11) |
| >35 years | 263 (34.65) |
| **Gender** |  |
| Male | 397 (52.30) |
| Female | 362 (47.69) |
| **Marital status** |  |
| Single | 327(43.08) |
| Married | 432 (56.91) |
| **Educational level** |  |
| Primary or less | 224(29.51) |
| Intermediate/Secondary | 284(37.41) |
| Higher education | 251(33.06) |
| **Occupation (Profession)** |  |
| Healthcare worker | 243(32.01) |
| Non-Healthcare worker | 516(67.98) |
| **Employment** |  |
| Unemployed | 567(74.70) |
| Employed | 192(25.29) |
| **Smoking status** |  |
| Yes | 223(29.38) |
| No | 525(70.61) |
| **History of chronic diseases** |  |
| Yes | 620(81.68) |
| No | 139(18.31) |
| **Infected with COVID19** |  |
| Yes | 417(54.94) |
| No | 342(45.05) |
| **Is the COVID-19 vaccine safe** |  |
| Yes | 521(68.77) |
| No | 237(31.22) |

According to the information presented in Figure 1, social media were considered a trustworthy source of information for COVID-19 vaccines by nearly 34% of the participants. Roughly 26%%, 21.8%, 15.6% and 12.48% of the participants placed their trust in Television, Healthcare provider, government and friends, respectively, as sources of information. On the other hand, only 3.43% of the participants trust sources of information got from scientific papers (Figure 1)

**Figure 1:**
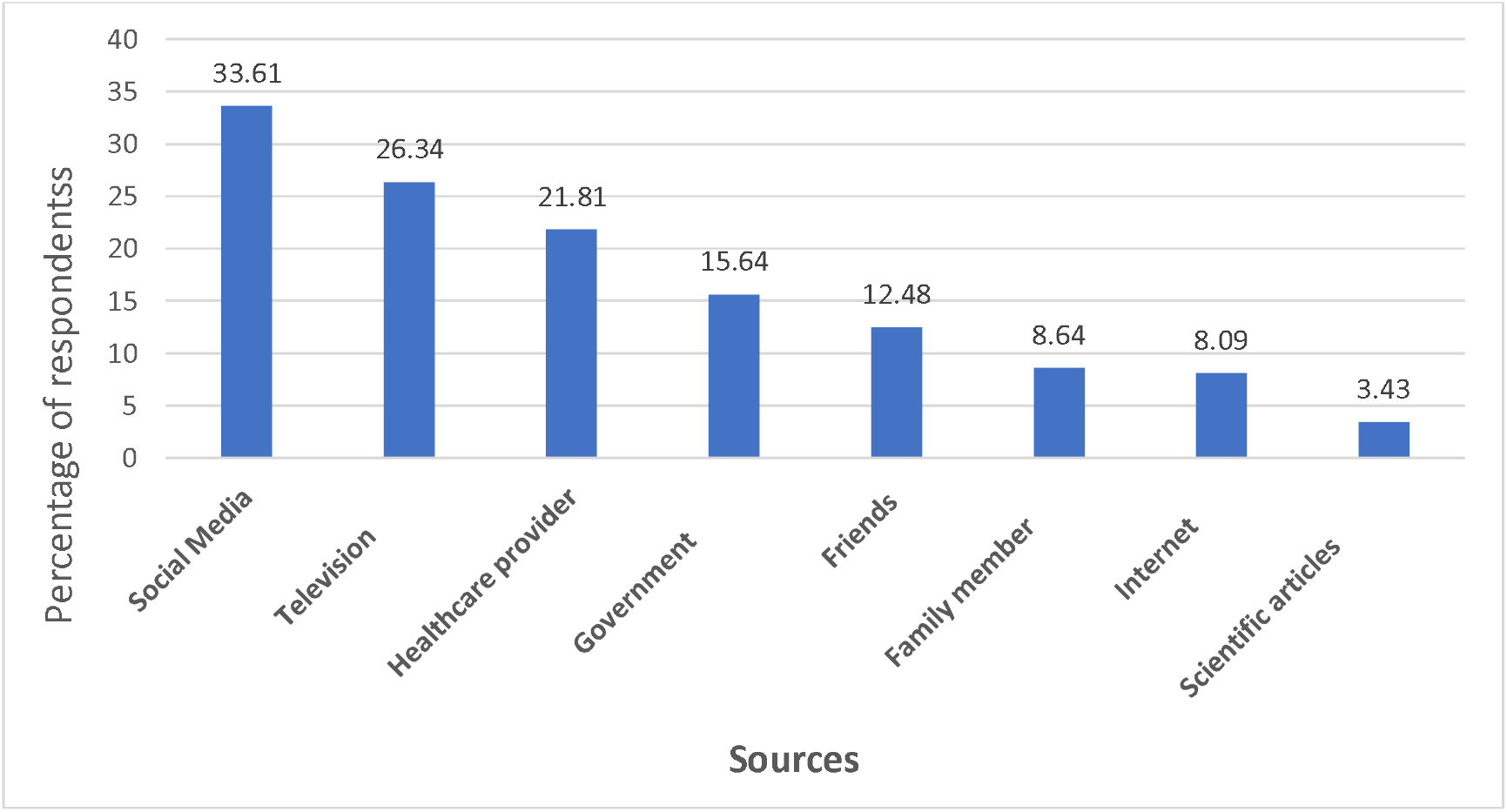
Source about COVID-19 vaccination.

### 3.2 Practice towards COVID19 vaccines

Figure 2 shows the reasons why individuals were unwilling to take the COVID-19 vaccine. Approximately 37% of the respondents reported that they did not have time to get vaccinated. Additionally, around 29% of participants indicated that they were uncertain about the safety of the vaccines, while about 18.6% reported that they believed the vaccine to be ineffective. Finally, approximately 14.5% of participants expressed fear of injections (Figure 2).

**Figure 2:**
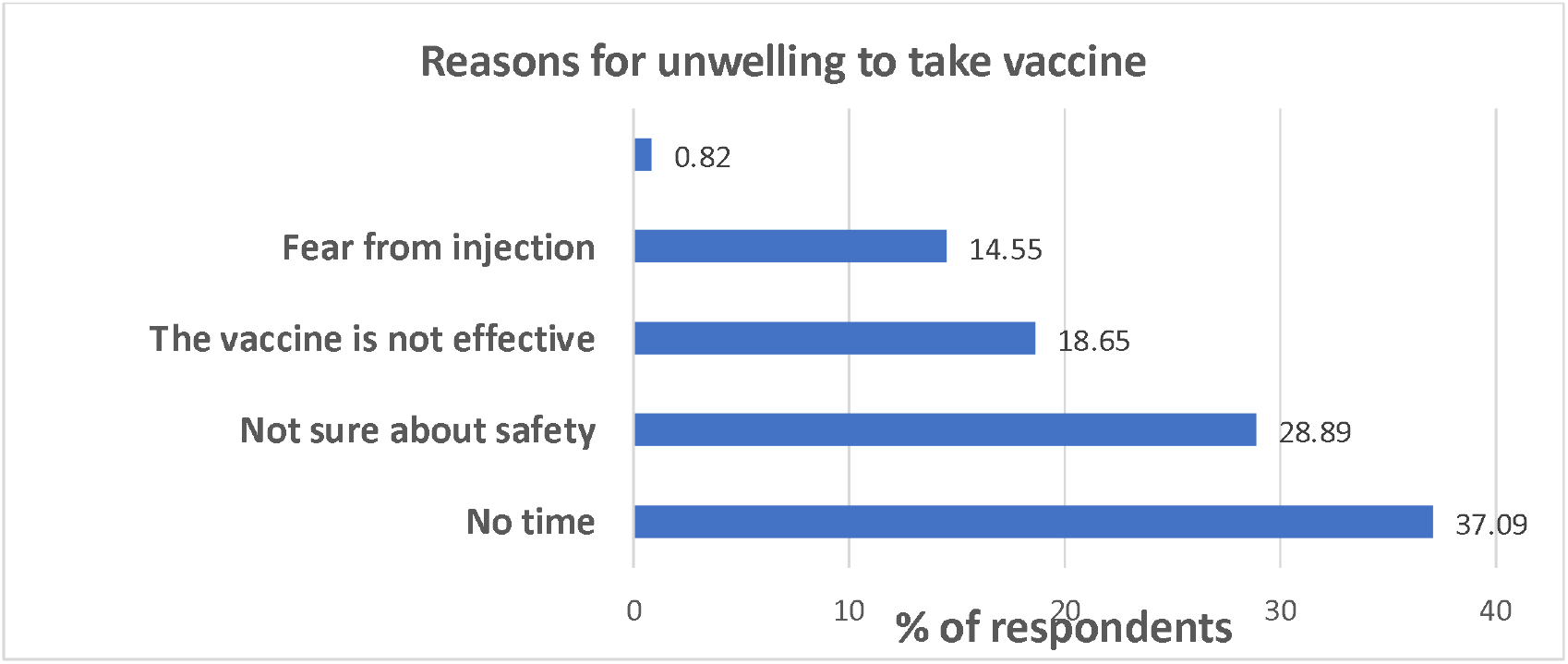
Reasons for unwillingness to take the COVID-19 vaccine.

### 3.3. Attitudes toward covid-19 vaccine

Almost three-quarters (74.04%) of the participants were strongly agreed that it is important to get a vaccine to protect the people from COVID-19. Besides, 59.55% of the participants reported that side effects will prevent them from taking a vaccine for the prevention of COVID-19 and that nearly two-thirds (66.53%) will refuse to take vaccine against COVID-19 once licensed. However, over half (55.59%) of the participants agreed that pharmaceutical companies will be able to develop safe and effective COVID-19 vaccines. Moreover, the majority (88.93%) of the respondents agreed that the government will make the vaccine available for all citizens for free. Around a quarter of all participants were disagreed regarding most attitudes as shown in Table 3. and that of them 5.14% disagreed that the government will make the vaccine available for all citizens for free.

**Table 3:** Attitudes toward covid-19 vaccine.

| Questions | Agree<br>No (%) | Neutral<br>No. (%) | Disagree<br>No. (%) |
| --- | --- | --- | --- |
| It is important to get a vaccine to protect the people from COVID-19 | 562 (74.04%) | 67 (8.83%) | 130 (17.13%) |
| Side effects will prevent me from taking a vaccine for the prevention of COVID-19 | 452 (59.55%) | 98 (12.91%) | 209 (27.54%) |
| Pharmaceutical companies are going to develop safe and effective COVID-19 vaccines | 422 (55.59%) | 172 (22.66%) | 165 (21.74%) |
| COVID-19 vaccines made in Europe or America are safer than those made in other world countries | 468 (61.66%) | 143 (18.84%) | 148 (19.49%) |
| Most people will refuse to take the COVID-19 vaccine | 505 (66.53%) | 106 (13.97%) | 148 (19.49%) |
| The government will make the vaccine available for all citizens for free | 675 (88.93%) | 45 (5.93%) | 39 (5.14%) |

### 3.4 Knowledge of COVID-19 vaccines

The majority of the participants (99.60%) had heard of COVID-19 vaccine, and (70.22%) of participant believed that the COVID-19 vaccine is safe with some side effects. Most participants (62.58%) thought that vaccines could protect them from COVID-19, and (73.25%) of participant believed that it is possible to get COVID-19 even after taking the COVID-19 vaccine. About (66.93%) of participant think that we can give the COVID-19 vaccine to a person with a history of COVID-19 and (65.48%) of them thought that the vaccine could not be given to a person who had symptoms of the disease at the time of vaccination. Beside (86.17%) of them knew about the side effects of the vaccine, (96.31%) of participants knew that the vaccines would be given in two doses or more. The overall knowledge related to the COVID-19 vaccine is shown in Table 4.

**Table 4:** Knowledge Regarding the COVID-19 Vaccine.

| **Questions** | **Yes (%)** | **No (%)** |
| --- | --- | --- |
| Have you heard about the COVID-19 vaccine | 756 (99.60) | 3 (0.395) |
| The COVID-19 vaccine is safe with some side effects | 533(70.22) | 226(29.78) |
| The COVID-19 vaccine protects from getting COVID-19 | 475(62.58) | 284(37.41) |
| It is not possible to get COVID-19 even after taking the COVID-19 vaccine | 203(26.75) | 556(73.25) |
| It is possible to give the COVID-19 vaccine to a person with a history of COVID-19 | 508(66.93) | 251(33.07) |
| It is not possible to give the COVID-19 vaccine to a person suffering from COVID-19 | 497(65.48) | 262(34.52) |
| Fever, slight swelling, and redness at the injection site are the side effects of the COVID-19 vaccine | 654(86.17) | 105(13.83) |
| The COVID-19 vaccine is given in 2 doses or more | 731(96.31) | 28(3.689) |

## Discussion

Coronavirus disease 2019 (COVID-19) is respiratory illness It was initially reported to the WHO on December 31, 2019 (1). The coronavirus (COVID-19) vaccines are safe and effective. They give the best protection against COVID-19. Everyone aged 5 and over can get a 1st and 2nd dose of the COVID-19 vaccine. People aged 5 and over who had a severely weakened immune system when they had their 1st or 2nd dose will be offered an additional primary dose (3rd dose) before any booster doses. Some people, including those aged 50 years or over, those at higher risk or who are pregnant, and frontline health and social care workers, will be offered a seasonal booster (autumn booster) (2)

During the COVID-19 pandemic, people used multiple information resources to gain knowledge and health information about the disease, including television, radio, newspapers, social media, friends, co-workers, healthcare providers, scientists, governments, etc (3)

In our study, more than half of the healthcare workers 64.0% were not vaccinated that is also high in Studies in Nigeria and Saudi Arabia reported intended vaccine uptake rates of 50.2% (4) and 50.52%, respectively (5) While vaccine acceptance rate was reported by a study in Iraq (61.7%) (6), which was higher than in two studies in the USA, where more than half of all healthcare workers were undecided and delayed the decision to be vaccinated (7,8) Low acceptance rates of vaccine were also reported among healthcare workers in Ghana (39.3%) (21), the Democratic Republic of Congo (27.7%)(22), Egypt (21%)(35), and Nepal (38.3%) (36) While highest vaccine acceptance reported from South and Southeast Asia, Studies in China and Vietnam reported intended vaccine uptakes of 76.63%(37) and 76.10%, respectively (38) These high rates were associated with good knowledge regarding the severity of COVID-19 and healthcare workers trust in the vaccines.

In our study the most common source trough people heard about covid-19 was social media (33.6%) then Television (26.3%), Healthcare provider (21.8%), Government (15.6%), and others were Family member, Friends, Internet and Scientific articles in lower ratios. While in study done in china in first line was social media (76.4%) then government (66.4%) and friends (54.5%) in lower rates (39). That was contrary to the study done in Jordan that health care provider was most used source for knowledge about covid vaccination (45.4%) then pharmaceutical companies (30.5%), government (27.1%) and others in lower rates (40). And also in contrast to a study done in Bangladesh that source of knowledge about COVID-19 vaccine was television (53%), social media (45%) and internet (38.7%), (41)that is same to a study done in Ethiopia television (20.5%), social media (17.3%) and internet (8.9%) (42) and also in a study done in Palestine the most common sources were social media (22%) and internet (18.7%) (43)

While unwilling to take COVID-19 vaccine was about 64.2% of the participant but in study in china only 24% (39) were unwilling to take vaccine and in study done in Oman 43% (44) were unwilling to take vaccine. And the most common reason reported in our study was not time about (37%) and second most common was not sure about safety (28.8%) others reported that vaccine is not effective, fear from injection and COVID is not a serious disease. It is contrary to a study done in Oman that only 0.3% reported no time, while 22.6% (44) not sure about safety same of us.

In our study’s findings illustrate that 99.60% of participants heard about the COVID-19 vaccine, and this was in line with similar studies conducted in oman demonstrating that knowledge significantly affects precautionary measures through the effectiveness of belief and had a direct effect on attitudes (45) The participants in our study who believed of the vaccine to be safe with some side effects were70.22% in contrast to study done in oman that only 17% believed in vaccine safety and effectivity and they considerably this low percentage due to the misinformation and rumors related to the vaccine, because the pandemic started in December 2019 and so much was unknown at its inception, misinformation evolved alongside facts about the disease, and this affected perceptions worldwide (45) And in another study done in Saudi Arabia 96.5% of participant believe that the vaccine is safe that is in similar line with our study (46) in study that conduct in Saudi Arabia the participant (66.8%) gotten covid even after being fully vaccinated, and only 42.1% believed that the COVID-19 vaccine prevents from spreading COVID-19. In our study 73.25% of participant believed that vaccine can’t prevent infection with covid again (47) healthcare personnels in Catania University Hospital (Italy) attitudes regarding mandatory vaccination for immunocompromised patient, newborn, and COPD to get some of necessary vaccines like (measles, varicella, and influenza vaccines) were increased and more supportive to get these vaccines during covid 19 pandemic, and (65%) of the staff they were intend to recommend SARS-CoV-2 vaccination to high-risk patients (48) In other side 62.58% of the participant believe that this vaccine can protect them from covid 19. In another study in Ethiopia about 93.9% of participant had known that there is second dose of vaccine (49)and this support our result that 96.31% of participant also known that there are second doses of vaccine.

The COVID-19 disease continues to spread with daily increase in the reported cases and deaths from countries overall the world (50)[26], while the world pins its hope on vaccines to prevent COVID-19. Before it is delivered to the market, COVID-19 vaccines need to be evaluated in humans for safety, efficacy and immunogenicity. This requires conduction of international clinical trials and recruitments of subjects from different countries to cover most of the population’s backgrounds. So knowledge, attitudes and practices (KAP) of the local population towards the COVID-19 vaccine is critical to understanding the epidemiological dynamics of disease control, and the effectiveness, compliance and success of the vaccination program (50). In this study our findings show that 74.04% of our sample population stated that it is important to get vaccinated to be protected against COVID-19, however, 61.53% agreed that most people would refuse to take the vaccine this is comparable with another cross-sectional study done in Iraq in which 61.40% reported an unwillingness to obtain a covid-19 vaccination (51). This disparity could be because of the thoughts they have regarding the vaccine’s adverse effects. It seems that specific details are not extremely important in the case of attitudes towards COVID-19 vaccine instead, general pro-vaccination principles, such as sufficient testing, safety, efficacy and being made in Europe and America should be focused on. Over the half (55.59%) of our sample population had confidence in pharmaceutical companies to develop safe and effective COVID-19 vaccines and this is comparable with another global cross-sectional study which showed that 57.60% of population had confidence in pharmaceutical companies to develop safe and effective COVID-19 vaccines. However, the source of the vaccine influences the perceived safety, as about two thirds (61.66%) of the participants in our study perceived that COVID-19 vaccines that were made in Europe or America were safer than those made in other world countries while in the previously mentioned study showed that about one-third (31.70%) of population perceived that COVID-19 vaccines that were made in Europe or America were safer than those made in other world countries (52).

## Conclusion

Our country is one of the highest countries for the refusal of COVID-19 vaccine with a rate of 64.2% influenced mainly by friends and social media. vaccination is one of the most effective interventions capable of controlling widespread infectious disease such as the current pandemic, but their outcome is based on the public’s trust in, and willingness to accept the vaccine. So it is imperative to increase people’s trust toward vaccine in order to improve its coverage and potentially control this pandemic.

## Data Availability

All data produced in the present work are contained in the manuscript

